# Burden and correlates of late diagnosis of cervical cancer among women attending care at a public health facility in Coastal Kenya

**DOI:** 10.64898/2026.07.14.26358118

**Authors:** Grace M. Mwango, Stevenson K. Chea

## Abstract

We aimed to assess the burden and correlates of late diagnosis of cervical cancer among women attending care at a public health facility in Coastal Kenya.

A retrospective cross-sectional design was used. Data extracted from medical records of women already diagnosed with cervical cancer at Kilifi County Referral Hospital (KCRH) were used. Logistic regression was used to assess independent predictors of late diagnosis of cervical cancer.

A total of 126 patients were included in this analysis. Out of the 126 participants, 67.5% (n=85) were 50 years and older with a mean age of 54 years. Among the 126 participants whose records were included in analysis, 73.0% (n=92; [95% CI: 64.3 – 80.5]) were diagnosed with late-stage cervical cancer. In multivariable analysis, none of the variables included in the model showed an association with the outcome.

In conclusion, we found a high burden of late diagnosis of cervical cancer in this setting. Targeted interventions are warranted to reverse this trend.

## Background

Cervical cancer remains a public health problem worldwide with sub Saharan Africa (SSA) reporting the highest incidence of approximately 20 cases per 100,000 women [1]. Cervical cancer is also one of the major causes of cancer mortality in SSA with an estimated 5 year survival of 33-35% [2, 3]. One of the effective strategies in cervical cancer control is screening [4]. Screening for cervical cancer facilitates early diagnosis thus timely intervention and better patient outcomes by detecting precancerous lesions before progression to invasive disease [4]. However, many SSA countries have low screening rates which may explain the high burden of late diagnosis [4]. It is not surprising therefore that SSA bears the greatest burden of cervical cancer mortality [4]. It is in recognition of this problem that the World Health Organization (WHO) coined the global strategy for elimination of cervical cancer as a public health problem [5]. The strategy sets the 90-70-90 target which aims to ensure 90% of girls are fully vaccinated against human papilloma virus by age 15 years; 70% of women are screened with a high-performance test by 35 years of age and again by 45 years of age and 90% of women diagnosed with cervical disease receive treatment [5].

Late diagnosis of cervical cancer has been studied in several studies from SSA ranging from 56.8% from an Ethiopian study [6] to 95.0% [7] in Kenya [2, 8-16]. A recent systematic review and meta analysis reported residence and education level as factors associated with late diagnosis [17]. A cross sectional study from Cote d’ Ivoire reported that being HIV infected, being uninsured and attending specialized facilities directly without being referred were significant predictors of late diagnosis of cervical cancer [8]. A 2024 study from Kenya identified high cost of travel to facility, advanced age and having anxiety over cost of cancer care as factors associated with increased odds of late diagnosis of cervical cancer [18]. A 2016 Ugandan study found that having few children and lack of money were associated with increased odds of late diagnosis of cervical cancer [13]. While studies have been done to assess correlates of late diagnosis of cervical cancer, the variation in findings suggests that findings may differ depending on context. Even with the observed variation in exposures assessed in the various studies, it is interesting that even sociodemographic indicators which are traditionally included in most analyses are also not performing the same way in the various studies. For example while age and socioeconomic status were significantly associated with late diagnosis of cervical cancer in a Kenyan study [18], none of the sociodemographic indicators included in analysis in a Ugandan study of 2024 was associated with late diagnosis of cervical cancer [9]. It makes sense therefore to study the prevalence of late diagnosis of cervical cancer and its drivers in the local context of Kilifi, Kenya. Identification of the burden of late diagnosis of cervical cancer and its drivers would guide targeted context-specific interventions to help fast track progress towards achieving the 90-70-90 goal. We aimed to assess the burden and correlates of late diagnosis of cervical cancer among women attending care at a public health facility in Coastal Kenya.

## Methods

### Study design and population

A retrospective cross-sectional design was used. Data extracted from medical records of women already diagnosed with cervical cancer at KCRH was used in this analysis.

### Study setting

In this study, existing data from medical records at KCRH were used. KCRH serves as the referral hospital in Kilifi County with a catchment population of about 1.5 million [19]. Cancer care at KCRH is provided as comprehensive cancer care [20]. Cervical cancer screening has been integrated with sexually transmitted infection care and family planning services [21] and is carried out in the Maternal and Child Health (MCH) department using a pap smear and via villi test[22]. WHO recommends that cervical cancer screening be done for all women starting at the age of 25 years and after every 3 to 5 years[4]. Women with a positive screen for cervical cancer undergo further investigations including a computed tomography (CT) scan to confirm the diagnosis and allow staging of the cancer. Staging is done using the International Federation of Gynecology and Obstetrics (FIGO) system [23]. In brief stage one means the cancer is limited to the cervix, in stage two the cancer cells have spread to tissues around the uterus or the upper two-thirds of the vagina. Stage three is when cancer involves the lower third of the vagina, may extend to the pelvic wall, lymph nodes and kidney functions may also be impaired, and finally, stage four is when the cancer has spread to the bladder lining, wall of rectum or other distant body parts [23]. Women confirmed to have cervical cancer get their clinical data relating to the diagnosis (including staging) included in their clinical files. Other data captured in the clinical files include socio demographic indicators including age and level of education. The women are then referred to Coast General Teaching and Referral Hospital, the largest referral hospital in Coastal Kenya, for cancer treatment while their clinical files are taken to the hospital archives for storage. According to records at KCRH, approximately 30 and 5 women are screened and confirmed to have cervical cancer every month respectively.

### Sources of data and measures

Hospital records (clinical files) of women who had been diagnosed with cervical cancer at KCRH from January 2022 up to end of 2024 were used. This was because these files were more accessible. The hospital records (hard copy patient files) were obtained from the archives where they are usually stored after diagnosis, staging and referral for treatment is done. Only files containing both socio-demographic and staging data were used. Data was manually extracted from the hard copy files and entered into an Excel sheet between 1^st^ July 2024 and 30^th^ August 2024. Authors had no access to information that could identify individual patients. Outcome data extracted was the stage of cervical cancer at diagnosis. Exposure data included age, level of education, residence, marital status and comorbidities. The outcome of interest in this study was late diagnosis of cervical cancer defined as having cervical cancer at stage 3 or 4 at diagnosis as per the FIGO staging system[23].

### Data analysis

Patient characteristics were described using frequencies and percentages and presented in a table. The number of records depicting women whose cervical cancer was at stage 3 or 4 at diagnosis was divided by the total records included in the analysis and multiplied by 100 to obtain the prevalence of late diagnosis. Binomial 95% confidence intervals [24] were presented. Then, univariable logistic regression [25]was conducted. Exposures with a p-value <0.2 were carried forward to the multivariable model. Finally, a multivariable logistic regression model was fitted using a stepwise model-building approach to determine the independent predictors of late diagnosis of cervical cancer. Odds ratios with their corresponding 95% confidence intervals as well as p values were presented in a table. All analysis was performed in Stata 15.0 (StataCorp. 2017. Stata Statistical Software: Release 15. College Station, TX: StataCorp LLC. 2019) and graphs generated using GraphPad Prism version 8.0.2 (GraphPad software, California).

### Ethical considerations

Ethical approval was granted by Pwani University Institutional Scientific and Ethics Review Committee (Ref: ISERC/BSc/058/2024). Informed consent was not directly sought since the current analysis used existing data from medical records.

## Results

### Participant characteristics

Between 2022 and 2024, 130 patients were diagnosed with cervical cancer at KCRH. Out of these, 4 records/files were missing thus files of 126 patients were included in this analysis. Out of the 126 participants, 67.5% (n=85) were 50 years and older with a mean age of 54. Further, majority, 91.3% (n=115) lived in rural areas, 81.7% (n=103) covered 0 to 50 kilometers to health facility, had completed primary education 47.6% (n=60) and were single 59.5% (n=75) (Table 1).

**Table 1:**
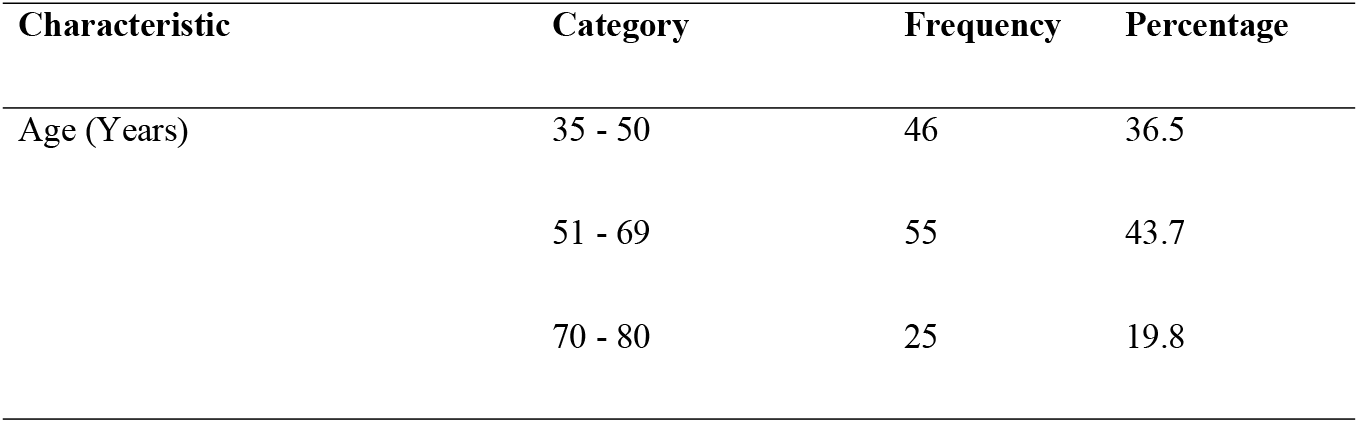
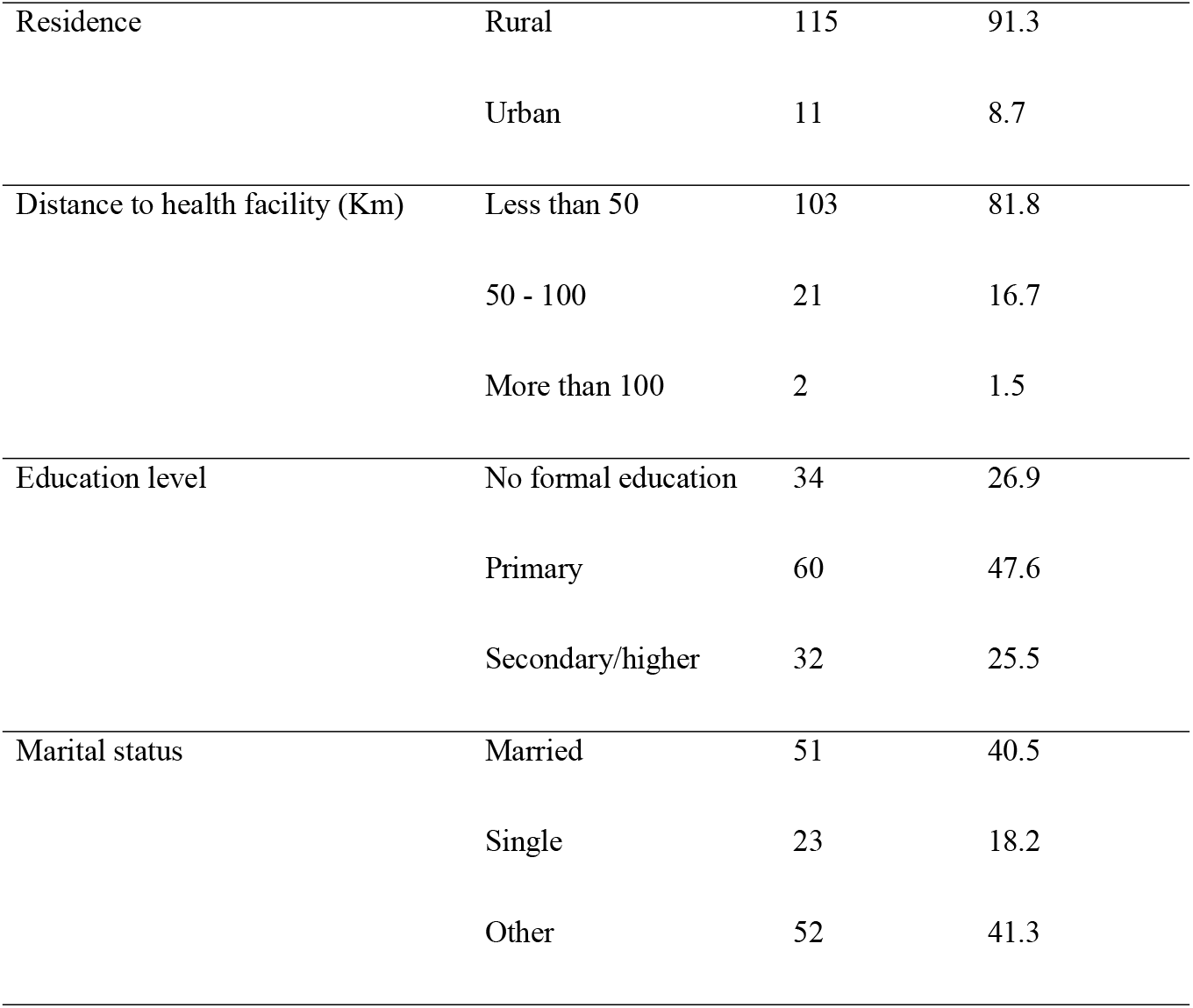
Sociodemographic characteristics of women diagnosed with cervical cancer at a public health facility in Coastal Kenya (N = 126)

| Characteristic | Category | Frequency | Percentage |
| --- | --- | --- | --- |
| Age (Years) | 35 - 50 | 46 | 36.5 |
|  | 51 - 69 | 55 | 43.7 |
|  | 70 - 80 | 25 | 19.8 |
| Residence | Rural | 115 | 91.3 |
|  | Urban | 11 | 8.7 |
| Distance to health facility (Km) | Less than 50 | 103 | 81.8 |
|  | 50 - 100 | 21 | 16.7 |
|  | More than 100 | 2 | 1.5 |
| Education level | No formal education | 34 | 26.9 |
|  | Primary | 60 | 47.6 |
|  | Secondary/higher | 32 | 25.5 |
| Marital status | Married | 51 | 40.5 |
|  | Single | 23 | 18.2 |
|  | Other | 52 | 41.3 |

### Burden of late diagnosis of cervical cancer

Among the 126 participants whose records were included in analysis, 73.0% (n=92; [95% CI: 64.3 – 80.5]) were diagnosed with late-stage cervical cancer. Further, majority of the women whose records were included in analysis had cervical cancer stage 3 at diagnosis 56.3% (n=71; [95% CI: 47.2 – 65.1]) (Fig 1).

**Fig 1:**
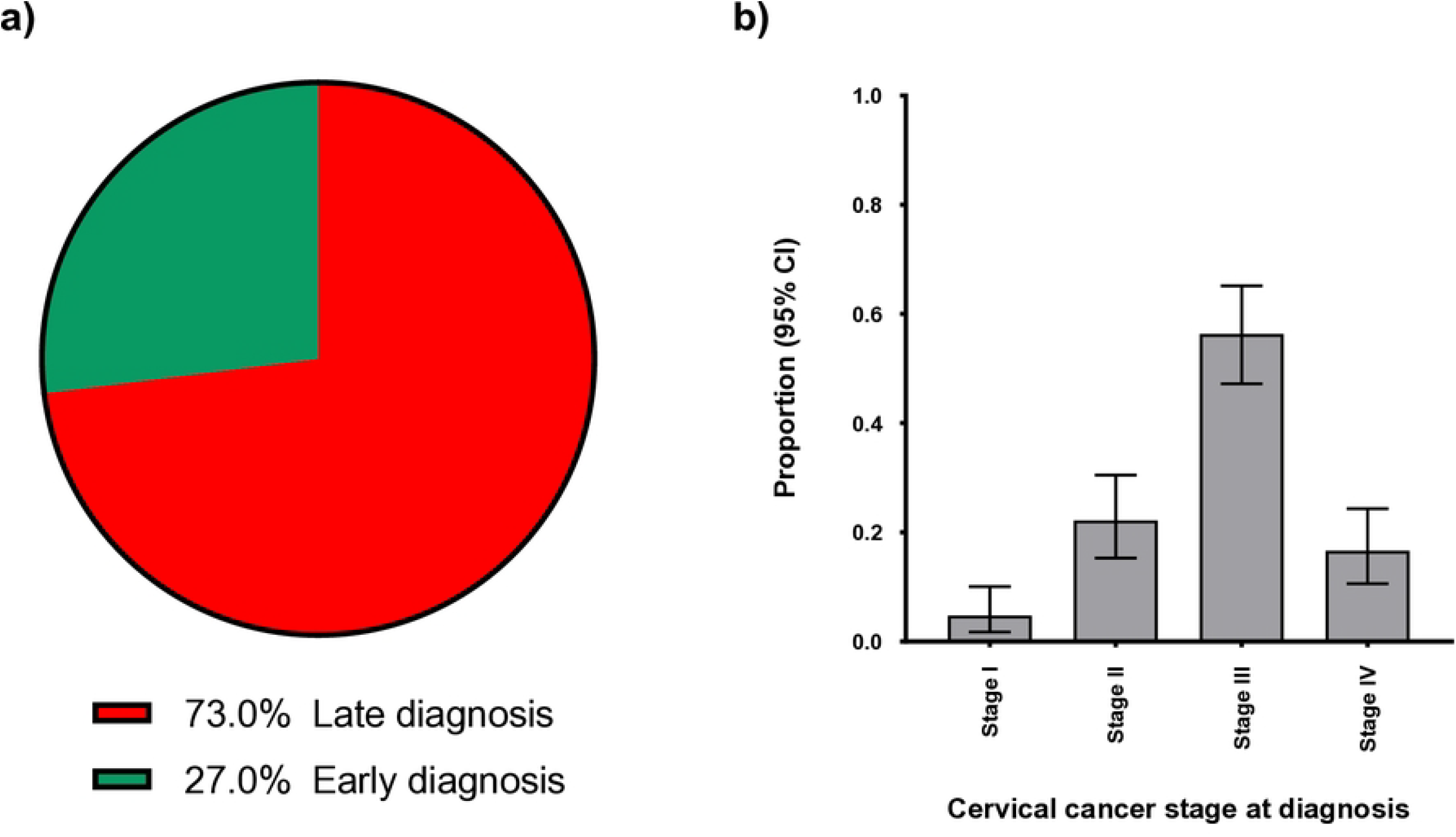
a) A pie chart depicting the proportion of women who were diagnosed with cervical cancer late (stage III or IV) and those diagnosed early (stage I or II) b) A bar graph showing distribution of women according to the stage of their cervical cancer at diagnosis as per the FIGO staging system

### Correlates of late diagnosis of cervical cancer

In univariable analysis, residence was associated with late diagnosis of cervical cancer. Specifically, women residing in rural areas were almost four fold as likely to have late diagnosis of cervical cancer compared to their urban counter parts (COR [95% CI]; p value) 3.7 [1.1 – 13.1]; p = 0.041]. In multivariable analysis, none of the variables included in the model showed an association with the outcome. Marital status attenuated residence towards the null (Table 2).

**Table 2:** Correlates of late diagnosis of cervical cancer among women diagnosed with cervical cancer at a public health facility in Coastal Kenya.

| Predictor | Category | Early<br>diagnosis<br>n[%] | Late<br>diagnosis<br>n[%] | Crude<br>OR<br>[95% CI] | P<br>value | Adjusted<br>OR<br>[95% CI] | P<br>value |
| --- | --- | --- | --- | --- | --- | --- | --- |
| Age [Years] | 35 - 50 | 13 [38.2] | 33 [35.8] | Ref | Ref | - | - |
|  | 51 - 69 | 15 [44.1] | 40 [43.5] | 1.1 [0.4 – 2.5] | 0.912 | - | - |
|  | 70 - 80 | 6 [17.7] | 19 [20.7] | 1.2 [0.4 -3.8] | 0.699 | - | - |
| Residence | Rural | 28 [82.4] | 87 [94.6] | 3.7 [1.1 – 13.1] | 0.041 | 3.3 [0.9 – 12.1] | 0.061 |
|  | Urban | 6 [17.6] | 5 [5.4] | Ref | Ref | Ref | Ref |
| Distance to health facility | <50 | 26 [76.5] | 77 [83.7] | Ref | Ref | - | - |
|  | ≥50 | 8 [23.5] | 15 [16.3] | 0.6 [0.2 – 1.6] | 0.354 | - | - |
| Education level | No formal education/primary | 23 [67.7] | 71 [77.2] | Ref | Ref | - | - |
|  | Secondary/higher | 11 [32.3] | 21 [22.8] | 0.6 [0.2 – 1.4] | 0.278 | - | - |
| Marital status | Married | 17 [50.0] | 34 [37.0] | Ref | Ref | Ref | Ref |
|  | Single/other | 17 [50.0] | 58 [63.0] | 1.7 [0.7 – 3.7] | 0.188 | 1.5 [0.6 – 3.4] | 0.300 |

## Discussion

In this study, almost 75 out of every 100 women attending care at a public health facility in coastal Kenya had a late diagnosis of cervical cancer. Sociodemographic indicators did not predict late diagnosis of cervical cancer.

We found a high burden of late diagnosis of cervical cancer in this setting. A previous study conducted in Cote d’ Ivoire that enrolled women diagnosed with cervical cancer between July 2018 and June 2019 reported that 71.5% of the women had advanced cervical cancer at diagnosis (late diagnosis) [8]. Another study from Kenya that enrolled cervical cancer patients seen at a referral hospital reported that at least 95% were diagnosed late [7]. A Ugandan cross-sectional study of women with histological diagnoses of invasive cervical cancer conducted between 2022 and 2023 at a referral hospital reported that 83.4% of the women enrolled had late stage cervical cancer at diagnosis [9]. These findings from previous studies suggest that late diagnosis of cervical cancer is not unique to the current study setting but reflects broader structural challenges in cancer control in resource limited settings and highlights the urgent need for strengthened cancer control strategies in settings like Kenya.

The high burden of late diagnosis in this setting may potentially be explained by barriers in accessing care and low education levels. Kilifi has been reported to have one of the highest rates of poverty in Kenya with overall poverty rate of about 46.0% compared to the national average of 36.1% [19]. Poverty has been shown to be a barrier in accessing care since people may not even have money to spend on transport to the health facility and other indirect costs associated with seeking health care [26-28]. Further, Kilifi has relatively low literacy levels compared to other regions in Kenya with a literacy level of approximately 68.0% [29]. Low literacy levels have been associated with a reduced understanding of medical advice including the importance of seeking care early, which can negatively impact health seeking behavior and outcomes [30, 31]. Additionally, the high burden of late diagnosis in this study reinforces concerns about missed opportunities for early detection. Evidence suggests that women in resource limited settings may have had prior contact with the health care system but are not comprehensively evaluated or referred to higher level facilities [32] thus delaying diagnosis. This finding of a high burden of late diagnosis of cervical cancer suggests that strengthening primary health care systems and improving diagnostic capacity of health care workers at lower level facilities could help reduce diagnostic delays.

Sociodemographic indicators did not predict late diagnosis of cervical cancer in this study. Previous studies have reported associations between sociodemographic indicators and late diagnosis of cervical cancer [13, 18]. This lack of an association may be due to the fact that only few exposures were included in analysis thus negatively impacting on the ability to detect an association. In the current study we analyzed data from hospital records. Hospital records are not designed for research thus may have short comings including a limited number of variables being collected [33]. Further, the lack of statistically significant predictors in the multivariable model may suggest limited statistical power due to the relatively small sample size. None the less, a previous study from Uganda that enrolled women with cervical cancer equally reported that sociodemographic indicators did not predict late diagnosis of cervical cancer [9]. Equally, a study from Nigeria reported that sociodemographic indicators were not associated with late diagnosis of cervical cancer [10].

An important strength of this study is that it is highlighting the burden of late stage diagnosis of cervical cancer in this setting which is a key determinant of treatment outcomes. However, this study has several limitations. First, the retrospective design may be subject to incomplete data and misclassification bias. Secondly, the study was conducted in one facility which may limit generalizability to other settings.

In conclusion, we found a high burden of late diagnosis of cervical cancer in this setting. Targeted context specific interventions are warranted to reverse this trend.

## Data Availability

Data relating to this study is freely available on figshare https://doi.org/10.6084/m9.figshare.32968310

